# Admission avoidance in UK healthcare: What works and what doesn’t? A hermeneutic review

**DOI:** 10.1101/2025.03.06.25323302

**Authors:** Cidalia Eusebio, Alison Leary

## Abstract

**Background:** Avoidable hospital admissions in the UK have been a focal area for healthcare policy and research, often with a focus on prevention. This is particularly important as hospitalisation can negatively impact patients, especially frail older people, leading to further risk of morbidity and mortality. To reduce costs and enhance patient care, the focus has shifted toward improving community interventions to prevent admissions. Actions to address this issue include the expansion of new types of care, such, for example, virtual wards. However, evidence is lacking on how current initiatives are improving care and which factors lead to emergency admissions which could be avoided in the UK. The aim of this study is to examine and synthesise the current evidence of admissions avoidance in the UK, including understanding how different populations, chronic conditions and social determinants might affect the most the risk of avoidable admissions. From a health services perspective the aim is to understand which types of care and community interventions prevent avoidable admissions.

**Design:** The published evidence on admission avoidance is voluminous and so a hermeneutic review was undertaken.

**Results:** 82 papers met the criteria. A dominant theme is the complexity of reducing emergency admissions, emphasising the impact of social determinants, particularly in deprived and older populations. Optimising service delivery, targeting care gaps, and improving care coordination are essential. Managing comorbidities, especially in conditions like ambulatory care-sensitive conditions such as COPD, diabetes, heart failure, cancer and preventing infections, can reduce admissions. Interventions vary in context, style and target with some, particularly those who provide care to high-risk patients and in deprived areas showing success in reducing emergency attendances and improving hospital resource use.

**Conclusion:** In conclusion, preventing avoidable admissions is complex due to the broad range of factors that influence risk of admission, and the multifaceted interventions required, as no single solution fits all neither interventions evaluations are consistent. This complexity should be considered when introducing new services. While shifting care towards the community can offer benefits, this review suggests a more integrated approach—one that bridges acute, hospital, and community care—is necessary to effectively reduce emergency admissions. Key target areas which might offer further opportunities are those populations who are elderly, experience poverty and social deprivation and which have high risk of comorbidities such as ambulatory care-sensitive conditions.

## Introduction

Avoidable hospital admissions in the UK have been a focal area for healthcare policy and research, aiming to reduce unnecessary healthcare costs and improve patient care. It is predicted 20% of admissions (1.5 - 2 million) could potentially be avoided every year with effective primary and community care interventions. [1] This is particularly important as hospitalisation can negatively impact patients, especially frail, leading to further risk of morbidity and mortality.

Some of the actions to tackle this problem have been expansion of new types of care, such, for example, integrated care models, virtual wards, social prescribing and new services or workforces such as frailty teams. [2] Although context-dependent, common areas for improvement have emerged in attempting to reduce admissions: lack of integration and communication in care transitions and the importance of physical enablers (supporting network and continuity of care). [3] With the proposed policy shift of care towards community it is important to ensure to understand the complexity of issues leading to avoidable admissions in order to target interventions and workforces that can provide the most cost-effective care.

However, evidence is lacking on what factors and how current initiatives in the community are avoiding admissions to emergency care. For example, in ambulatory care sensitive conditions (ACSC), variations in emergency avoidance may arise from sociocultural factors, unmet needs, or non-healthcare influences like community, employment, and housing. [4] Although there are studies looking at changes in activity which are then adopted, there is varying success as it is not possible to see what currently works for who, why and in what circumstances - in such a complex system it is important to understand which measures are effective for which population groups and which workforces need to be deployed or developed to achieve the best outcomes.[5] It is important to know which specific interventions work best for distinct patient groups and in what circumstances they are most effective. By understanding which admission avoidance strategies are most effective, the review can provide evidence-based guidance to inform the investment in scalable interventions in primary and community care but also to identify gaps in existing studies, where more robust research is needed to produce reliable knowledge on long term outcomes and cost-effectiveness.

The issue of avoidable emergency admissions presents a complex challenge within healthcare, requiring an approach going beyond traditional evidence synthesis. This hermeneutic review aims to address this complexity by employing an iterative critical interpretive approach, connecting evidence across diverse sources to foster a more interpretative understanding of the underlying factors and interrelationships.

A hermeneutic review is an interpretative process focussing on developing a deep understanding of existing literature. It is iterative in nature, involving continuous dialogue between the researcher and the texts, where interpretation and critical reflection evolve with each engagement. [6] It is also helpful where diverse methodologies are used and allows a wider range of studies to be synthesised. This process involves engaging in a cyclical process known as the hermeneutic circle. It starts with literature searching, followed by critical reading and interpretation, leading to a refined understanding. As information is gained, literature is revisited, reinterpreting earlier texts considering new understanding, gradually building a coherent and nuanced perspective on the research problem. [6]

The aim is to understand the factors which may lead to emergency admissions as well exploring the interventions evaluated across the diverse UK healthcare setting as well looking at their cost-effectiveness and outcomes. Additionally, the review prioritises research addressing the role of social determinants of health, care models, or community care in preventing hospital admissions.

## Methods

The hermeneutic review was chosen to reflect the complexity of the issues and to gain a broad understanding of not only the causes of avoidable admissions, population factors, social determinants and well as factors related to access to healthcare interventions in the community and their impact in reducing avoidable admissions. Compared with a systematic review, this allows a wider perspective and interpretation with a wider search and iterative analysis.

A search was made of the following databases (MEDLINE, PubMed, Google Scholar, EMABASE, CINAHL and Cochrane library) with the following search terms:(“avoidable admission*” OR “preventable hospital*” OR “unplanned admission*” OR “emergency admission*”) (“chronic disease*” OR “multi-morbidity” OR “elderly” OR “older adult*” OR “high-risk population*”) (“social determinant*” OR “socioeconomic factor*” OR “healthcare access” OR “rural-urban disparity” OR “deprivation” OR “housing”) (“community care” OR “integrated care” OR “home-based care” OR “virtual ward*” OR “telehealth” OR “remote monitoring” OR “primary care intervention*”).

The review included studies conducted in the United Kingdom, published in English, and focused on avoidable or preventable hospital admissions. Studies examine populations aged 18 and older and published within the last 10 years (2014-2024) to ensure alignment with the current healthcare policy landscape.

Studies conducted outside the UK or focused solely on populations under 18 years old were excluded. Research focused solely on non-avoidable admissions or acute care that does not address the prevention of admissions will be excluded to maintain the review’s relevance to avoidable admissions.

The search generated 1786 publications. After removal of duplicates and evidence which did not meet our inclusion criteria, 82 publications remain for a hermeneutic review. Results were collected into an excel spreadsheet. Each study’s design, date, population, sample size, key findings, research gaps, and needs were documented. Each study was categorised by primary themes identified through the research, including data-driven preventive interventions, acute and community-level interventions, GP access, social determinants, quality and outcomes frameworks (QOF) and audits, qualitative patient research, prediction models, and the financial impacts of admissions. Each study was further refined into sub-themes based on specific diseases (e.g., heart failure, COPD) or target populations (e.g., older people).

Through multiple hermeneutic cycles a re-evaluation of the studies was completed, enabling understanding of factors and interventions contributing to avoidable admissions. Collaboration with experts further contextualised findings, highlighting key core themes and their interrelationships.

## Results

The themes found in the review of the literature reflect the complexity of the issue both the challenges that society faces in terms of healthcare service provision and the influence of social determinants of health, as well as potential interventions and their associated costs (figure 1). While the themes were partly shaped by the search strategy, the specific characteristics of each theme and the relationships between them were uncovered through the iterative process of hermeneutic cycles

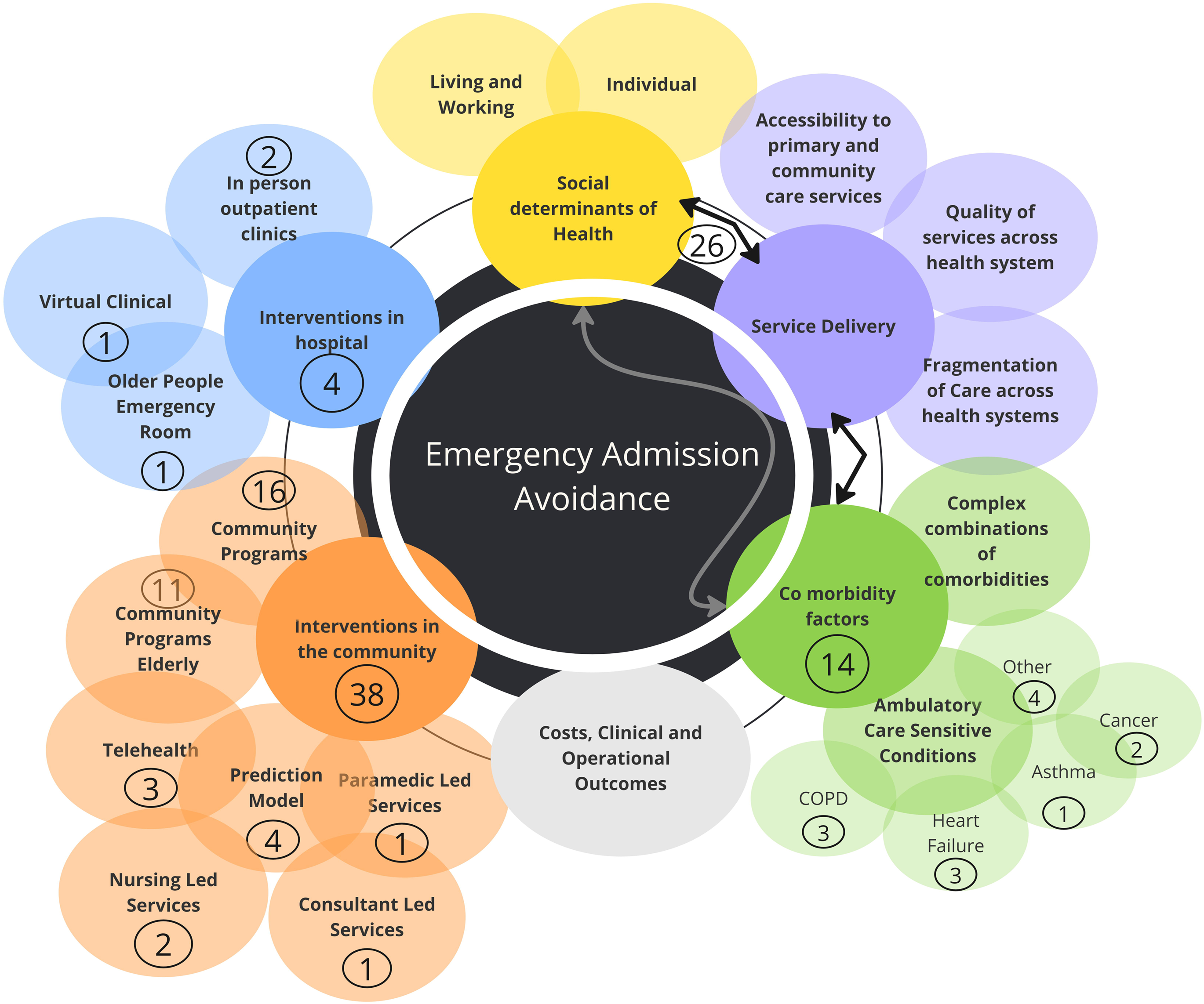

### The impact of social determinants of Health in Emergency Admissions

Social determinants of health had a greater emphasis through studies, both at the broad population level as well as those focusing on specific cohorts of patients. Living and working conditions as well as individual health factors seem to predominate the study findings.

Living alone, especially in the older population, emerged as a significant factor, with several studies highlighting its role in increased emergency visits. [7] Other contributing factors included deprivation, social isolation, frailty, lack of social support, and cohabitation with frail individuals.[7] Geographic variation also played a role, with differences observed between rural and urban areas. [8]

Home adaptations, particularly for older adults, and the use of living aids were found to reduce hospital admissions. [9] From an individual health perspective, key contributors to emergency admissions included age (particularly for patients over 65), gender (depending on comorbidities), certain comorbidities, life-threatening events, and an elevated risk of falls. [9] Those who were more engaged in planned care tended to experience more elective admissions and utilised non-GP primary care services more frequently, reducing the pressure on emergency departments. [10]

### The impact of service delivery in Emergency Admissions

Access and delivery of healthcare services significantly impact on emergency admissions, especially due to care quality and fragmentation between community, primary, acute care factors. Care quality issues were not limited to primary care; they extend across the entire health system.

From a care quality perspective, factors such as poor hospital discharge processes, lack of comprehensive care plan and delays in transitioning patients back into the community were found to contribute to emergency readmissions.[11] Additionally, inadequate patient and carers education also played a role. [12]. The lack of expertise in managing complex patients, coupled with perhaps poor continuity of care and low-quality care in settings like care homes, had an impact in emergency admissions. [13]

Fragmentation between acute services, primary care, and community care often exacerbates patient outcomes and unplanned emergency admissions, particularly for older, frail adults, those on chemotherapy and heart failure patients. [12] Additionally, in primary and community care, a lack of resources and the significant variability in service provision, particularly between urban and rural regions, were also highlighted. [14] Limited access to general practitioners (GPs) and geriatricians (within emergency departments), further influenced unplanned emergency admissions, especially in conditions such as heart failure, diabetes and urinary tract infections/pyelonephritis. [15]

The absence of preventative care support systems, insufficient care/support to manage symptoms at home, limited access to palliative care, poor care coordination, and a general lack of integrated care strategies and alternatives to hospital admissions all contribute to increased emergency service utilisation.[12] Furthermore, weak leadership within primary and community care, combined with the absence of coordinated care strategies, creates an environment where emergency admissions are more likely to occur. [15]

### The impact of Specific Comorbidities on Emergency Admissions

In analysing emergency attendances, the review highlighted two key areas where avoidable admissions are prevalent. They are often accompanied by outcome disparities particularly when social determinants are present related to social economic deprivation. The first area is related to patients with complex combinations of comorbidities, such as heart failure, chronic kidney disease (CKD), and hypertension, which are associated with a high risk of readmission to hospital. [16] The second area involves specific conditions, particularly ambulatory care-sensitive conditions (ACSC) like Chronic Obstructive Pulmonary Disease (COPD), diabetes, Urinary tract infections, asthma and heart failure. [8]

Several studies highlighted a lack of accessibility to community and primary care services, especially related to disease management, prevention and palliative care. For example, individuals with intellectual disabilities and cognitive impairments were found to be at higher risk of readmission, often due to mismanagement of their ACSC, which could have been avoided with appropriate care and prevention. [17] Depression and dementia also contributed to higher readmission rates, particularly among older adults. [18,19] Among cancer patients, specialist nurses reduced emergency admissions by proactively managing care. [20] Additionally, in colorectal cancer patients, most readmissions were linked to chemotherapy complications or insufficient symptom management in the community, rather than post-surgical issues, leading to reliance on emergency care. [21]

In terms of costs, prevalent combinations, such as hypertension and diabetes, contributed more to overall system costs, while more complex combinations (with 6+ conditions) generated higher per-patient costs. Preventable emergency admissions were most associated with combinations including chronic heart failure, chronic kidney disease, and hypertension. [16]

### Interventions which support reduction in Emergency Admissions

The review of literature examining interventions aimed at reducing emergency admissions revealed a variety of approaches, ranging from hospital-based to community-based interventions/programs, as well as data-driven interventions using machine learning and artificial intelligence to support prevention strategies. Most evaluated interventions focused on short-term solutions, rather than long-term strategies in specific geographic areas, with limited cost-effectiveness evaluation. While many did not significantly reduce emergency admissions, some improved clinical and operational outcomes, such as, for example, shorter hospital stays and better disease management.

Hospital-based interventions included in-person lounge clinics, virtual clinics, and specialised emergency departments. In-person lounge clinics focused on specific comorbidities such as heart failure and limb ischemia, with both examples showing promising outcomes. For heart failure, the introduction of a diuretic lounge led to a 13% reduction in emergency admissions, with an estimated annual cost savings of £315.000/annum and £2900 per admission avoided. Although long-term hospital stays increased for these patients, short-term admissions were reduced, indicating effective outpatient management. [22] In the case of limb ischemia, there were no significant differences in readmission rates, but the intervention saved 441 bed days, reduced hospital stays from an average of 17 to 3 days, and improved clinical outcomes— patients were six times more likely to avoid surgical intervention and 2.5 times less likely to require amputation. [23]

Virtual clinics, nurse-led, such as those used for renal stones, also showed positive results. These virtual follow-ups reduced the cost per appointment by 93%, resulting in a cost saving of £12,000, with only 5% of patients requiring emergency admission. [24] Telephone advice provided by hospital centres by specialist nurses for cancer patients in the community, although not evaluated quantitatively, has emphasised their importance in providing advice on symptom management at home but also be a point of contact for triage and assessment for those who require admissions. [20]

A study evaluated hospitals that implemented specialised emergency departments for older adults, found mixed results.[25] While shorter waiting times, quicker clinical assessments, and a higher likelihood of patients being discharged home were gained, there was no significant reduction in overall hospital admissions or differences in mortality or re attendance rates compared to standard emergency departments. [25]

Looking at community-based interventions aimed at reducing emergency admissions, there was a diverse array of programmes. These included telehealth interventions for conditions such as kidney transplants, COPD, and diabetes, clinician lead services, and various disease-specific community programs targeting conditions like COPD, diabetes, neuromuscular disease, stroke, and interventions specifically designed for older adults.

Telehealth interventions showed some benefits, such as minor cost savings and improvements in medication management, emotional support, and patient self-management. However, most studies did not demonstrate significant reductions in emergency admissions.

Some interventions looked at clinician lead services. Paramedic-led services, such as those for diabetes, were more successful, especially in deprived areas where on-site treatment for conditions like hyperglycemia helped prevent unnecessary hospital admissions. [26] In consultant-led community services for COPD, although the overall reduction in emergency admissions was not significant, these services had a meaningful impact on specific groups, particularly in deprived areas, where COPD-related emergency admissions were significantly reduced. [27]

Community programs for diabetes showed variability in outcomes, but in those focused on achieving a high number of quality clinical targets, significant reductions in emergency admissions were observed. [28] For example, achieving HbA1c and cholesterol targets reduced the risk of all emergency admissions by 9%, and cardiovascular admissions by 7-12%.[28] Annual reviews with updated care plans showed to have benefits in reducing hospital admissions in ACSC (23%) in severe mental health patients and 26% in patients with disability. [29]

Programmes focusing on integrated care had mixed results. A programme providing multidisciplinary meetings, discharge liaison and social prescribing had positive results with over 800 avoided monthly referrals to emergency admissions, especially among patients aged 75 years old and a decrease in emergency bed days by 50% leading savings of 3.8 million annually and improving staff satisfaction.[30] Although these outcomes did not translate in other studies with similar interventions which only saw temporary effects in reduction of emergency admission. [31]

Interventions targeting other diseases, such as neuro motor disease (NMD), reduced emergency readmissions from 25% to 12%, shortened hospital stays, and improved mortality rates. This success was attributed to a higher proportion of NMD patients being managed by specialised neuromuscular centres, coordinated multidisciplinary care, stronger links, and improved education for both patients and non-specialist acute teams through a network/partnership approach involving charitable organisations. [32] Approaches to stroke care were also tested with a mobile stroke unit demonstrated a 25% reduction in unnecessary hospital admissions and significantly improved thrombosis times by bypassing emergency departments and transferring patients directly to stroke units, resulting in higher thrombosis rates compared to the UK average. [33]

Interventions aimed at older adults varied widely, with most failing to sustain long-term reductions in emergency admissions. Interventions, particularly aimed at older adults, such as holistic assessments and integrated care programs, were found to be cost-neutral or not cost effective, with no significant reduction in healthcare expenditure observed. [34]

Medication management also emerged as a significant issue, with 84% of older adults in the community having potentially inappropriate prescriptions, which were associated with higher risks of hospital admission and mortality. Additionally, polypharmacy (defined as five or more medications) was linked to an increased likelihood of inappropriate prescribing and prescription omissions.[35] Finally, prediction models tailored to specific diseases, which incorporated complex variables, showed promise in forecasting hospital admissions. However, challenges remain in improving not only the types of predictors such as social determinants but also the accuracy of 30-day readmission, predictions and ensuring generalizability across different studies and populations. [36]

## Discussion

This hermeneutic review highlights the complexity of factors involved in reducing and avoiding emergency admissions, highlighting the need for a system-wide, integrated approach. These three key areas, social determinants, comorbidities and service delivery, are not distinct but interdependent of each other and therefore should be considered when designing new interventions to prevent emergency admissions.

Social determinants of health, especially among deprived populations, older individuals with socioeconomic challenges, and those without adequate care coordination significantly contribute to emergency admissions. Targeting these high-risk groups is crucial to reduce emergency attendance as previous predictive studies have found.[3] While the strategic shift towards community care and addressing social determinants is promising as well also seen in some interventions analysis, austerity and government disinvestment over the past decades have deepened health inequalities, as evidenced in the Marmot Review. [37]

The COVID-19 pandemic and the cost-of-living crisis have only exacerbated these inequalities. Research by the Health Foundation shows that a 60-year-old in the poorest areas of England has the same level of diagnosed illness as a 76-year-old woman or 70-year-old if man in the wealthiest areas. [38] The growing health inequalities highlighted by Marmot reinforce the view that the real causes behind health failures are social, with a graded relationship between deprivation and health. [37]

Equally important is improving the delivery service by identifying and addressing gaps in care quality. Implementing targeted interventions to strengthen these areas is essential. An integrated care strategy that enhances care coordination across regions, hospital to community care and ensures appropriate resource allocation is critical for improving access to community-based care. Although this is not often straightforward.

Integrated care strategies critically depend on well-coordinated care, with numerous studies reporting coordination as a key issue. [39] This is not only crucial for avoiding admissions but also prominently featured in coroner’s reports as a factor in preventing future deaths. Issues like deficits in skill and knowledge, delayed or uncoordinated care, and communication barriers highlight that uncoordinated care can be fatal—not only can it prevent admission avoidance, but it may directly compromise patient safety. [40]

In the community, coordination is typically provided by district nurses or specialist nurses within specific fields, such as cancer care. However, two significant challenges currently affect these efforts: the reduction of district nurses by 40%, and an overwhelming caseload—with over 30% of teams managing more than 400 patients per nurse, and around 60% of teams frequently deferring or delaying care. This strain is a systemic issue with managing individual caseloads effectively. [41]

Nurses are essential to proactive case management; they don’t just deliver physical care but also provide emotional, cognitive, and organisational support—types of labour that can often remain hidden or under-acknowledged due to limited evidence on nursing-lead interventions and outcomes in district nursing care. [42] Still, as seen in this review, some nursing interventions seem promising in terms of outcomes. [43]

Another example, looked at proactive case management interventions by multiple sclerosis specialist nurses have shown significant reductions in care utilisation, decreasing from an average of 2,700 to just 198 bed days per year. [44] This demonstrates the clear role of nurses in not only direct patient care but also in organising and coordinating complex support across multiple health services. Nonetheless, a workforce shortage limits this potential, and it’s clear that effective coordinated care requires access to skilled multidisciplinary professionals who can tailor support to specific patient needs.

Implementing a large-scale, sustainable, and effective strategy is often complex, requiring a clear understanding of factors supporting service innovation, adoption, sustainability, and scaling. [45] Some identified by literature include strong leadership and management aligned with a clear and compelling vision, broad stakeholder engagement, dedicated and ongoing resources, and effective communication across organisations. [46] Additionally, adapting to local contexts, providing continuous monitoring and timely feedback, and demonstrating cost-effectiveness through strong evaluation are essential for sustaining and scaling integrated care initiatives. [46]

Addressing comorbidities is another key factor, as different conditions require tailored interventions. Special attention should be given to ambulatory care-sensitive conditions in patients with higher socioeconomic deprivation. Focus should be given particularly to COPD, diabetes, heart failure. Other conditions like learning disabilities, cognitive impairments, depression, dementia, cancer (particularly in managing chemotherapy-related conditions) are often associated with increased emergency attendance as well. Complex comorbidities, such as the combination of heart failure, chronic kidney disease (CKD), and hypertension, further contribute to elevated emergency admission rates, especially when compounded by infections and as well as the social determinants of health.

The literature presents a broad array of interventions, but the diversity of terminology used, regional disparities, varying study designs, and limited representativeness make it challenging to draw broad conclusions. Nevertheless, some targeted interventions have demonstrated success in reducing both emergency admissions and hospital stays, thereby enhancing resource management. Improving care coordination, medication management and compliance with national clinical guidelines for specific conditions has improved outcomes and reduced admissions, but stronger outcomes were achieved in most deprived populations. Previous literature reviews have found case management associated with a greater adherence to treatment guidelines and higher patient satisfaction, however, did not translate into better use of hospital resources. [47]

Despite the expansion of virtual wards across the NHS, their monitoring and evaluation in relation to avoidable admission and their cost-effectiveness remains underexplored across health systems with limited evidence on their clinical effectiveness. [48] Two Randomised Control Trials (RCT) have found that virtual physician care was non inferior, but in-home-care was still needed for 25% of patients. [49] On contrary hospital-at-home programs, (at least a visit of a nurse or doctor) present significant evidence. Previous systematic reviews reveal that hospital-at-home interventions can help patients with specific chronic diseases (e.g. heart failure and COPD) avoid admissions, reduce the length of stay, improve patient satisfaction as well as improve clinical effectiveness and cost-effectiveness compared to inpatient care. [50]

### Limitations

This review offers a broad, health system-level analysis of factors and interventions to avoid emergency admissions in the United Kingdom but lacks the depth necessary for implementing more targeted, context-specific approaches nor compares other healthcare systems who can offer different approaches. There are few studies that focus on specific diseases, and the interventions examined are often geographically limited and short-term, with robust cost evaluations rarely conducted. Moreover, the absence of randomised, longitudinal studies weakens the overall evidence base, emphasising the need for more rigorous research to assess the long-term impact of interventions aimed at reducing emergency admissions.

### Areas for Future Research

Future studies should adopt a system-wide, longitudinal approach to evaluating interventions aimed at reducing emergency admissions. Research should focus not only on population-level impacts and medication management but also on disease-specific interventions to assess their cost-effectiveness.

Ambulatory care-sensitive conditions and comorbidities, particularly in deprived populations, demand more research. Rigorous studies, including randomised controlled trials, are needed to evaluate the long-term effectiveness of interventions in reducing emergency admissions. Further research should focus on different types of virtual wards and their effectiveness in specific populations as well as the evaluation of hospital-at-home programs.

Additionally, predictive models, though in their early stages, show potential for supporting long-term community interventions and enhancing cost-effectiveness in care. More research is required to explore how predictive tools can be applied to better support community-based care.

### Implications for policy

While health policy increasingly promotes a push towards community-based care, as the NHS Long Term plan proposes, a strategic approach is necessary to ensure effectiveness. Hospital-supported interventions for high-risk, complex patients may also play a crucial role in preventing avoidable admissions. Broader changes to community and acute services may be required to improve access not only to clinical decision-makers but also to specialised care, particularly in areas such as cancer and heart failure. This includes ensuring the capacity to deliver care and access necessary treatments.

Enabling multidisciplinary specialist care within communities, without relying solely on general practitioners, could bridge gaps in care. Tailored, disease-specific approaches should be adapted to regional needs, supported by adequate funding and resources to ensure access.

Greater access to palliative care is also critical, particularly for patients with chronic diseases and cancer, who could significantly benefit from better symptom management and specialised care.

## Conclusion

This analysis highlights the complexity of preventing avoidable admissions due to the broad range of factors influencing risk and the need for multifaceted interventions. No single solution fits all scenarios, and a system-wide, integrated approach is essential. A key focus should be on social determinants of health, particularly among at-risk populations, such as those facing deprivation, elderly individuals with socioeconomic challenges, and those without adequate care support.

Equally important is addressing gaps in service delivery, particularly in care quality and coordination between hospitals and community care. Targeted interventions that enhance care coordination across different services are crucial in reducing emergency admissions. Comorbidities, especially those Ambulatory sensitive Care conditions show opportunities for improvement.

## Data Availability

All data produced in the present work are contained in the manuscript.

## REFERENCES

1 Torjesen I. Almost 1.5m emergency hospital admissions could have been avoided last year. BMJ. 2018;361:k2542. doi: 10.1136/bmj.k2542

2 NHS England » FRAIL strategy. https://www.england.nhs.uk/long-read/frail-strategy/ (accessed 28 October 2024)

3 Kayyali R, Funnell G, Odeh B, et al. Investigating the characteristics and needs of frequently admitting hospital patients: a cross-sectional study in the UK. BMJ Open. 2020;10:e035522. doi: 10.1136/bmjopen-2019-035522

4 Hodgson K, Deeny SR, Steventon A. Ambulatory care-sensitive conditions: their potential uses and limitations. BMJ Qual Saf. 2019;28:429–33. doi: 10.1136/bmjqs-2018-008820

5 Webb WA, Mitchell T, Snelling P, et al. Life’s hard and then you die: the end-of-life priorities of people experiencing homelessness in the UK. Int J Palliat Nurs. 2020;26:120–32. doi: 10.12968/ijpn.2020.26.3.120

6 Boell SK, Cecez-Kecmanovic D. A Hermeneutic Approach for Conducting Literature Reviews and Literature Searches. CAIS. 2014;34. doi: 10.17705/1CAIS.03412

7 Lloyd T, Crellin E, Brine RJ, et al. Association between household context and emergency hospital use in older people: a retrospective cohort study on indicators for people living alone or living with somebody with frailty, developed from routine healthcare data in England. BMJ Open. 2022;12:e059371. doi: 10.1136/bmjopen-2021-059371

8 Alsallakh MA, Rodgers SE, Lyons RA, et al. Association of socioeconomic deprivation with asthma care, outcomes, and deaths in Wales: A 5-year national linked primary and secondary care cohort study. PLoS Med. 2021;18:e1003497. doi: 10.1371/journal.pmed.1003497

9 Dew R, Wilkes S. Attitudes, perceptions, and behaviours associated with hospital admission avoidance: a qualitative study of high-risk patients in primary care. Br J Gen Pract. 2018;68:e460–8. doi: 10.3399/bjgp18X697493

10 Bu F, Fancourt D. How is patient activation related to healthcare service utilisation? Evidence from electronic patient records in England. BMC Health Serv Res. 2021;21:1196. doi: 10.1186/s12913-021-07115-7

11 Kamwa V, Knight T, Atkin C, et al. Acute frailty services: results of a national day of care survey. BMC Geriatr. 2024;24:608. doi: 10.1186/s12877-024-05075-1

12 Glover G, Williams R, Oyinlola J. An observational cohort study of numbers and causes of preventable general hospital admissions in people with and without intellectual disabilities in England. J Intellect Disabil Res. 2020;64:331–44. doi: 10.1111/jir.12722

13 Sherlaw-Johnson C, Smith P, Bardsley M. Continuous monitoring of emergency admissions of older care home residents to hospital. Age Ageing. 2016;45:71–7. doi: 10.1093/ageing/afv158

14 Moreo K, BSN R-B, BHSA C. Community and Case Management Interventions to Address Social Determinants of Health in Infectious Disease. Prof Case Manag. 2021;26:4–10. doi: 10.1097/NCM.0000000000000453

15 Wilson A, Baker R, Bankart J, et al. Understanding variation in unplanned admissions of people aged 85 and over: a systems-based approach. BMJ Open. 2019;9:e026405. doi: 10.1136/bmjopen-2018-026405

16 Stokes J, Guthrie B, Mercer SW, et al. Multimorbidity combinations, costs of hospital care and potentially preventable emergency admissions in England: A cohort study. PLoS Med. 2021;18:e1003514. doi: 10.1371/journal.pmed.1003514

17 Hosking FJ, Carey IM, DeWilde S, et al. Preventable Emergency Hospital Admissions Among Adults With Intellectual Disability in England. Ann Fam Med. 2017;15:462–70. doi: 10.1370/afm.2104

18 Guthrie EA, Dickens C, Blakemore A, et al. Depression predicts future emergency hospital admissions in primary care patients with chronic physical illness. J Psychosom Res. 2016;82:54–61. doi: 10.1016/j.jpsychores.2014.10.002

19 Sommerlad A, Perera G, Mueller C, et al. Hospitalisation of people with dementia: evidence from English electronic health records from 2008 to 2016. Eur J Epidemiol. 2019;34:567–77. doi: 10.1007/s10654-019-00481-x

20 Stewart S, Robertson C, Kennedy S, et al. Personalized infection prevention and control: identifying patients at risk of healthcare-associated infection. J Hosp Infect. 2021;114:32–42. doi: 10.1016/j.jhin.2021.03.032

21 Ang CW, Seretis C, Wanigasooriya K, et al. The most frequent cause of 90-day unplanned hospital readmission following colorectal cancer resection is chemotherapy complications. Colorectal Dis. 2015;17:779–86. doi: 10.1111/codi.12945

22 Ioannou A, Browne T, Jordan S, et al. Diuretic lounge and the impact on hospital admissions for treatment of decompensated heart failure. QJM. 2020;113:651–6. doi: 10.1093/qjmed/hcaa114

23 Khan A, Hughes M, Ting M, et al. A “hot clinic” for cold limbs: the benefit of urgent clinics for patients with critical limb ischaemia. Ann R Coll Surg Engl. 2020;102:412–7. doi: 10.1308/rcsann.2020.0068

24 Hughes T, Pietropaolo A, Archer M, et al. Lessons Learnt (Clinical Outcomes and Cost Savings) from Virtual Stone Clinic and Their Application in the Era Post-COVID-19: Prospective Outcomes over a 6-Year Period from a University Teaching Hospital. J Endourol. 2021;35:200–5. doi: 10.1089/end.2020.0708

25 Meechan C, Navaneetharaja N, Bailey S, et al. EVALUATION OF THE FIRST OLDER PEOPLE’S EMERGENCY DEPARTMENT IN ENGLAND - A RETROSPECTIVE COHORT STUDY. J Emerg Med. 2023;65:e50–9. doi: 10.1016/j.jemermed.2023.04.003

26 van Woerden H, Bucholc M, Clubbs Coldron B, et al. Factors influencing hospital conveyance following ambulance attendance for people with diabetes: A retrospective observational study. Diabet Med. 2021;38:e14384. doi: 10.1111/dme.14384

27 Saini P, Rose T, Downing J, et al. Impact of community-based chronic obstructive pulmonary disease service, a multidisciplinary intervention in an area of high deprivation: a longitudinal matched controlled study. BMJ Open. 2020;10:e032931. doi: 10.1136/bmjopen-2019-032931

28 Chandok R, FRCGP F, GP Principal D, et al. An update to collaborating for improving diabetes care in Ealing, London: investment, integration and innovation, years 2011-2020. 2022;39:23–9. doi: 10.1002/pdi.2376

29 Ride J, Kasteridis P, Gutacker N, et al. Do care plans and annual reviews of physical health influence unplanned hospital utilisation for people with serious mental illness? Analysis of linked longitudinal primary and secondary healthcare records in England. BMJ Open. 2018;8:e023135. doi: 10.1136/bmjopen-2018-023135

30 Withers K, Pardy K, Topham L, et al. A long way from Frome: improving connections between patients, local services and communities to reduce emergency admissions. BMC Prim Care. 2024;25:307. doi: 10.1186/s12875-024-02557-4

31 Exley J, Abel GA, Fernandez J-L, et al. Impact of the Southwark and Lambeth Integrated Care Older People’s Programme on hospital utilisation and costs: controlled time series and cost-consequence analysis. BMJ Open. 2019;9:e024220. doi: 10.1136/bmjopen-2018-024220

32 Scalco RS, Quinlivan RM, Nastasi L, et al. Improving specialised care for neuromuscular patients reduces the frequency of preventable emergency hospital admissions. Neuromuscul Disord. 2020;30:173–9. doi: 10.1016/j.nmd.2019.11.013

33 Grunwald IQ, Phillips DJ, Sexby D, et al. Mobile Stroke Unit in the UK Healthcare System: Avoidance of Unnecessary Accident and Emergency Admissions. Cerebrovasc Dis. 2020;49:388–95. doi: 10.1159/000508910

34 Moffatt S, Wildman J, Pollard TM, et al. Impact of a social prescribing intervention in North East England on adults with type 2 diabetes: the SPRING_NE multimethod study. Public Health Res (Southampt). 2023;11:1–185. doi: 10.3310/AQXC8219

35 Counter D, Millar JWT, McLay JS. Hospital readmissions, mortality and potentially inappropriate prescribing: a retrospective study of older adults discharged from hospital. Br J Clin Pharmacol. 2018;84:1757–63. doi: 10.1111/bcp.13607

36 Garcia-Arce A, Rico F, Zayas-Castro JL. Comparison of Machine Learning Algorithms for the Prediction of Preventable Hospital Readmissions. J Healthc Qual. 2018;40:129–38. doi: 10.1097/JHQ.0000000000000080

37 Marmot M. Health equity in England: the Marmot review 10 years on. BMJ. 2020;368:m693. doi: 10.1136/bmj.m693

38 The Health Foundation. Quantifying health inequalities in England. 2022. https://www.health.org.uk/news-and-comment/charts-and-infographics/quantifying-health-inequalities (accessed 18 October 2024)

39 Lhussier M, Dalkin S, Hetherington R. Community care for severely frail older people: Developing explanations of how, why and for whom it works. Int J Older People Nurs. 2019;14:e12217. doi: 10.1111/opn.12217

40 Leary A, Bushe D, Oldman C, et al. A thematic analysis of the prevention of future deaths reports in healthcare from HM coroners in England and Wales 2016–2019. Journal of Patient Safety and Risk Management. 2021;26:14–21. doi: 10.1177/2516043521992651

41 District Nursing Today. The Queen’s Nursing Institute. https://qni.org.uk/resources/district-nursing-today-the-view-of-district-nurse-team-leaders-in-the-uk/ (accessed 1 November 2024)

42 Jackson J, Anderson JE, Maben J. What is nursing work? A meta-narrative review and integrated framework. International Journal of Nursing Studies. 2021;122:103944. doi: 10.1016/j.ijnurstu.2021.103944

43 Stewart I, Leary A, Khakwani A, et al. Do working practices of cancer nurse specialists improve clinical outcomes? Retrospective cohort analysis from the English National Lung Cancer Audit. Int J Nurs Stud. 2021;118:103718. doi: 10.1016/j.ijnurstu.2020.103718

44 Leary A, Quinn D, Bowen A. Impact of Proactive Case Management by Multiple Sclerosis Specialist Nurses on Use of Unscheduled Care and Emergency Presentation in Multiple Sclerosis: A Case Study. International Journal of MS Care. 2015;17:159. doi: 10.7224/1537-2073.2014-011

45 World Health Organization. How do we ensure that innovation in health service delivery and organization is implemented, sustained and spread? 2018.

46 Greenhalgh T, Robert G, Macfarlane F, et al. Diffusion of Innovations in Service Organizations: Systematic Review and Recommendations. The Milbank Quarterly. 2004;82:581–629. doi: 10.1111/j.0887-378X.2004.00325.x

47 Buja A, Francesconi P, Bellini I, et al. Health and health service usage outcomes of case management for patients with long-term conditions: a review of reviews. Primary Health Care Research & Development. 2020;21:e26. doi: 10.1017/S1463423620000080

48 Norman G, Bennett P, Vardy ERLC. Virtual wards: a rapid evidence synthesis and implications for the care of older people. Age and Ageing. 2023;52:afac319. doi: 10.1093/ageing/afac319

49 Levine DM, Paz M, Burke K, et al. Remote vs In-home Physician Visits for Hospital-Level Care at Home: A Randomized Clinical Trial. JAMA Network Open. 2022;5:e2229067. doi: 10.1001/jamanetworkopen.2022.29067

50 Pandit JA, Pawelek JB, Leff B, et al. The hospital at home in the USA: current status and future prospects. npj Digit Med. 2024;7:1–7. doi: 10.1038/s41746-024-01040-9

